# Beyond Consensus: Human Ground Truth, Expert Disagreement, and Clinical Relevance in Pathology AI

**DOI:** 10.64898/2026.11.18.26368395

**Authors:** Makoto Kawamoto, Kris Lami, Frederico Gaia, Sosuke Ishijima, Shintaro Sato, Rosane Duarte Achcar, Amna R. Almutrafi, Richard Attanoos, Jaroslaw Augustyniak, Mary Beth Beasley, Sabina Berezowska, Luka Brcic, Alberto Cavazza, Wei-Chin Chang, Andrew Churg, Wendy A. Cooper, Alexandre Todorovic Fabro, Akira Hebisawa, Chutima Jiragawasan, Izidor Kern, Yuka Kitamura, Kristine E. Konopka, Shinobu Kunugi, Brandon T. Larsen, Tereza Losmanova, Mai Matsumura, Alberto Marchevsky, Jijgee Munkhdelger, Ellen Caroline Toledo do Nascimento, Koji Okudera, Noriah Binti Othman, Ryoko Saito-Koyama, Tomoe Sawazumi, Frank Schneider, Yuri Tachibana, Angela Takano, Fabio Tavora, Yukio Tsushima, Lee Fong Wan, Joanne L. Wright, Masayo Yoshimura, Wataru Uegami, Anil Parwani, Andrey Bychkov, Junya Fukuoka

## Abstract

Human consensus is often treated as ground truth in clinical artificial intelligence (AI), yet greater agreement does not necessarily ensure greater clinical relevance. We investigated how expert panel size, composition, and consensus formation influence the reproducibility and clinical relevance of reference standards across three independent pathology cohorts evaluated by international expert pathologists: 50 transbronchial lung cryobiopsy cases from patients with interstitial lung disease assessed by 27 pathologists, 95 resected lung adenocarcinomas evaluated by 11 pathologists, and 596 images of pulmonary granulomatous lesions evaluated by 12 pathologists.

Larger panels produced more stable collective judgments but did not improve associations with disease progression or survival in the first two cohorts, instead converging toward consensus classifications with weaker outcome associations. Groups of pathologists identified solely by their diagnostic behavior generated consensus classifications with differing clinical relevance. In lung adenocarcinoma, examination of cases graded differently by the all-rater consensus and a distinct pathologist cluster revealed specific morphological patterns potentially contributing to these differences, suggesting that expert disagreement may contain clinically informative signals. In the granuloma cohort, a modified Delphi process increased agreement, with Krippendorff’s α rising from 0.351 to 0.539, while disease–morphology associations changed only modestly.

These findings demonstrate that reproducibility and clinical relevance are distinct properties of expert-derived reference standards. Expert disagreement should therefore not be regarded solely as noise to be eliminated, but also as a potential source of clinically meaningful information. Identifying and independently validating outcome-associated morphological features may enable refinement of existing consensus frameworks, supporting the development of more clinically relevant, task-specific ground truth for pathology AI.

## Introduction

Clinical artificial intelligence increasingly depends on human-generated reference labels (1). In pathology, radiology, and other image-based disciplines, models are commonly trained or evaluated against labels produced by individual specialists, majority voting, adjudication, or structured expert consensus (1, 2). When no definitive reference test exists, however, such labels are not directly observed truths but constructed reference standards (3,4).

Consensus methodology has established important principles for expert selection, independence of initial judgments, structured feedback, and transparent decision-making (5). Diagnostic-methodology studies have likewise shown substantial heterogeneity in how expert panels are assembled and how their opinions are converted into reference standards. Bertens et al. identified major variation in expert-panel methodology and reporting (4), and a recent systematic scoping review confirmed persistent heterogeneity in panel composition, consensus procedures, and handling of uncertainty (2). Waggoner et al. similarly emphasized that consensus methodology itself requires explicit design choices (5).

A less explored question is what happens when highly experienced subspecialty experts still disagree.

Such disagreement may reflect more than random error. Experts can differ systematically in feature recognition, diagnostic thresholds, and the weighting of competing evidence (6). Collective-intelligence research has shown that majority opinion is not inevitably optimal when specialized information is concentrated within a minority, and that the method of aggregating judgments can materially affect collective performance (7). Rather than treating such disagreement solely as noise, examining the morphological features underlying divergent judgments may reveal clinically informative signals that could otherwise be overlooked during consensus formation.

This issue is particularly important for AI. Label noise affects medical-image models (8), but the reference-label generation process itself can also influence downstream performance (9). Annotation quality depends on labeling instructions (10), and studies in radiology have shown that low-agreement “gold” labels can depend strongly on which experts constitute the panel (11). Approaches that explicitly model multiple annotators rather than simply collapsing their opinions have therefore gained interest (12).

Pathology provides a particularly relevant setting because substantial disagreement can persist even among experts. Recent melanoma research demonstrated clinically meaningful diagnostic discordance among expert pathologists and questioned how reference standards for AI should be established (13).

Our previous lung adenocarcinoma studies provided a related example (6, 14). We identified internally coherent expert groups based on diagnostic behavior and subsequently used alternative expert-derived consensus standards to train separate deep-learning models. These studies demonstrated that structured variation in expert judgment can propagate into computational systems rather than disappearing during AI development.

Here, we examined three dimensions of expert reference-standard architecture: panel size, expert composition, and consensus formation. Across three independent pathology cohorts, we evaluated their relationships with two distinct properties of the resulting reference standard: consensus stability or agreement, and external clinical relevance.

We deliberately focused on diagnostic tasks in which interobserver variability is clinically and methodologically relevant (15). Our objective was not to identify a universally optimal panel or rank individual experts, but to determine whether reference-standard architecture changes the information retained within expert consensus and whether clinically informative disagreement should be understood before it is eliminated. We also explored whether discordant expert judgments could reveal morphological features that might inform future refinement of consensus-based reference standards.

## Methods

### Study design and expert panels

We conducted a retrospective multi-cohort study examining three components of expert-derived reference-standard architecture: panel size, expert composition, and consensus formation (Figure 1).

**Figure 1.**
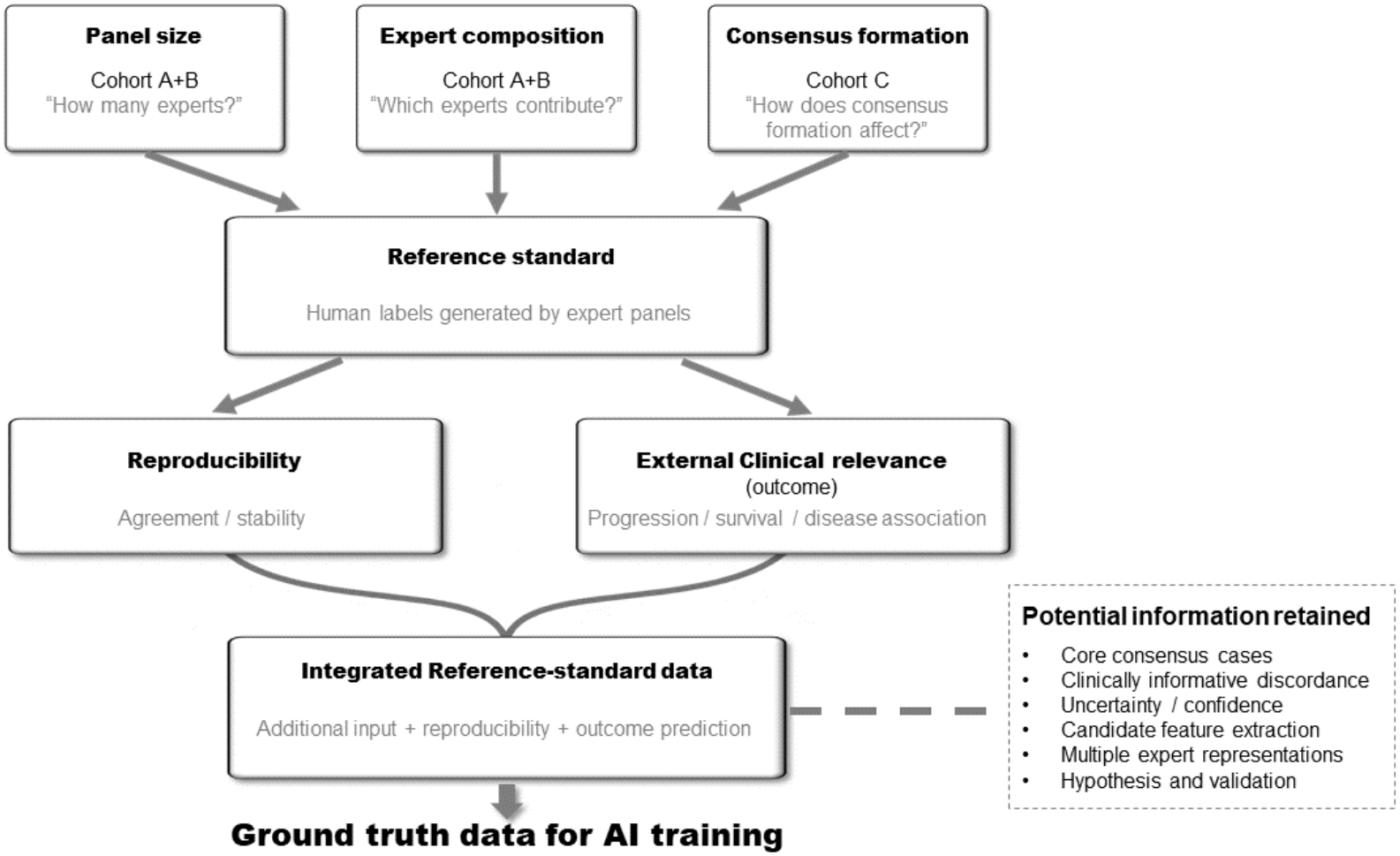
Conceptual framework for expert-derived reference-standard design in clinical AI. Reference standards were evaluated as products of three expert-panel design factors: panel size, expert composition, and consensus formation. Panel size and expert composition were examined in Cohorts A and B, and consensus formation in Cohort C. Reference-standard quality was assessed separately in terms of reproducibility and external clinical relevance. The framework further proposes that these complementary dimensions, together with structured expert disagreement, may be integrated to generate more informative training and validation targets for downstream AI. This integration is presented as a conceptual extension of the study findings rather than as a validated reference-standard construction method.

The three reader studies were originally established as independent international projects intended to generate expert-derived reference data for downstream AI research. Panel sizes were not predetermined. Participating pulmonary pathologists were those who agreed to contribute to the respective project and completed the required assessments.

Cohort A comprised 27 experts from 15 countries, Cohort B included 11 pathologists from five countries, and Cohort C included 12 experts from seven countries. The mean years of experience in pulmonary pathology were 22, 14, and 20 years, respectively. All participants in Cohorts A and C held academic positions as professors. Cohort B included four practicing pathologists without professor titles who were also actively involved in lung cancer pathology. Thirteen experts in Cohort A, three in Cohort B, and seven in Cohort C had contributed to the development of guidelines or classification systems at the international, national, or regional level, including WHO classifications (Table 1).

**Table 1.**
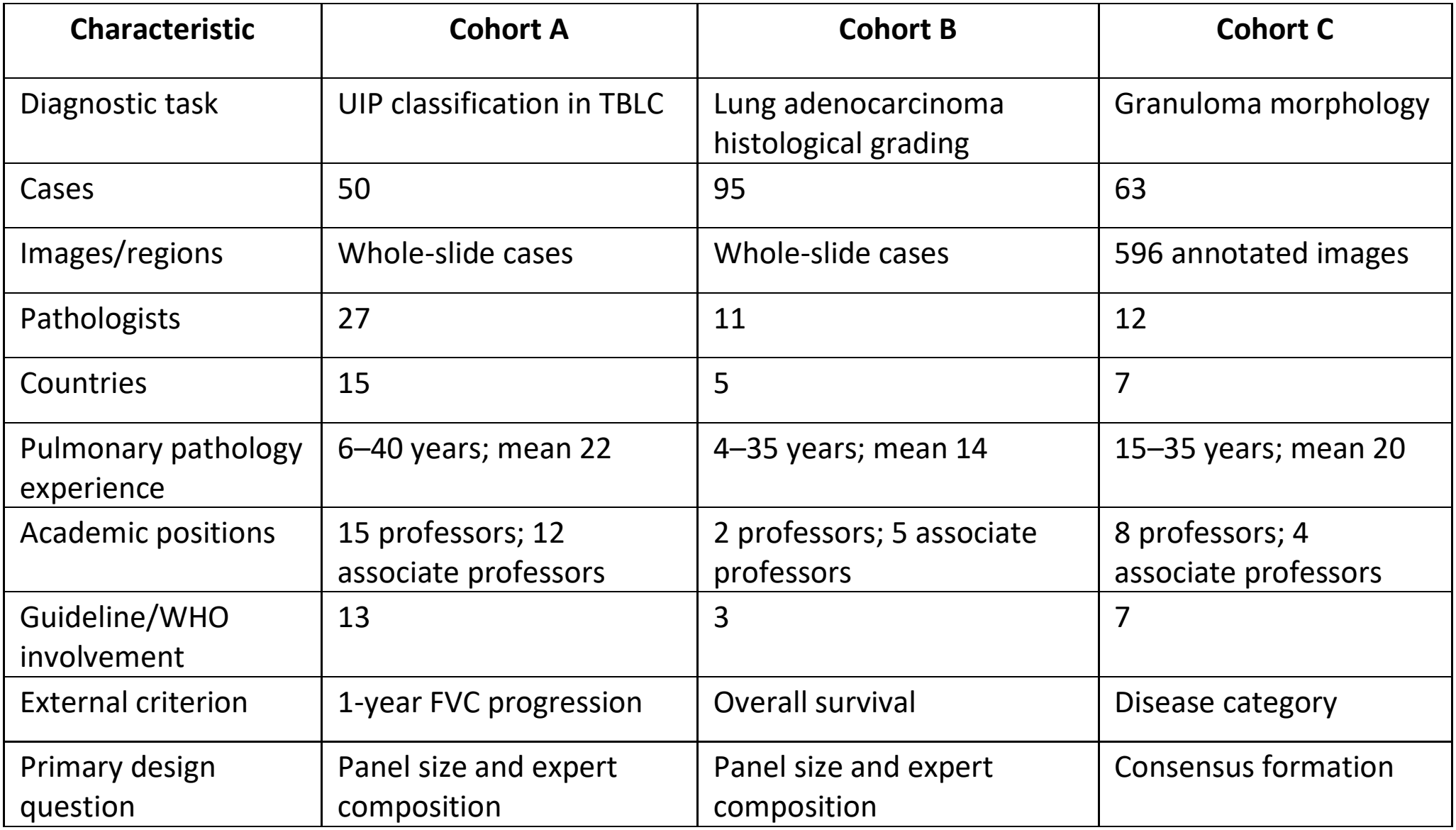
Study cohorts and expert panels.

| **Characteristic** | **Cohort A** | **Cohort B** | **Cohort C** |
|---|---|---|---|
| Diagnostic task | UIP classification in TBLC | Lung adenocarcinoma histological grading | Granuloma morphology |
| Cases | 50 | 95 | 63 |
| Images/regions | Whole-slide cases | Whole-slide cases | 596 annotated images |
| Pathologists | 27 | 11 | 12 |
| Countries | 15 | 5 | 7 |
| Pulmonary pathology experience | 6–40 years; mean 22 | 4–35 years; mean 14 | 15–35 years; mean 20 |
| Academic positions | 15 professors; 12 associate professors | 2 professors; 5 associate professors | 8 professors; 4 associate professors |
| Guideline/WHO involvement | 13 | 3 | 7 |
| External criterion | 1-year FVC progression | Overall survival | Disease category |
| Primary design question | Panel size and expert composition | Panel size and expert composition | Consensus formation |

Initial assessments were performed independently without access to other experts’ responses.

For this study, reproducibility refers to agreement or stability of expert-derived classifications, whereas external clinical relevance refers to association with a clinical or disease-related external criteria such as patients’ survival and disease progression.

External clinical relevance was not considered equivalent to morphological truth.

Ethics approval was obtained from Nagasaki University (IRB: 17051510-4).

### Cohort A: interstitial lung disease

Cohort A consisted of 50 patients who underwent transbronchial lung cryobiopsy for evaluation of interstitial lung disease.

Twenty-seven pulmonary pathology experts independently classified usual interstitial pneumonia (UIP) morphology using four categories: not UIP, presence of UIP as a minor component, UIP as a major component, and UIP. For binary analyses, UIP as a major component and UIP were grouped as UIP-group.

Disease progression was defined as an absolute decline of at least 5 percentage points in percent-predicted forced vital capacity (FVC) within 1 year, either from the time of diagnosis or from the first available pulmonary function test. Follow-up data were available for 49 patients.

### Cohort B: lung adenocarcinoma

Cohort B consisted of 95 surgically resected lung adenocarcinomas independently evaluated by 11 pulmonary pathologists.

Histologic grading was performed according to the IASLC grading system for invasive pulmonary adenocarcinoma (16). Tumors were classified as Grade 1 when lepidic predominant with <20% high-grade patterns, Grade 2 when acinar or papillary predominant with <20% high-grade patterns, and Grade 3 when ≥20% high-grade patterns were present, irrespective of the predominant pattern. High-grade patterns included solid, micropapillary, and complex glandular patterns.

Overall survival was used as the external clinical endpoint.

### Cohort C: pulmonary granulomas

Cohort C consisted of 63 patients with granulomatous or granuloma-like pulmonary lesions and final diagnoses of hypersensitivity pneumonitis, idiopathic pulmonary fibrosis, sarcoidosis, or granulomatosis with polyangiitis.

A total of 596 annotated images of candidate granulomatous lesions were evaluated. Twelve experts completed both assessment rounds and were included in paired pre/post-Delphi analyses (17).

Morphological assessments were consolidated into four categories: palisading granuloma, well-formed granuloma, poorly formed granuloma, and non-granuloma.

After Round 1, anonymized aggregate classification results were returned to participants. The same experts then independently reassessed the images in a modified Delphi process.

### Panel-size analysis

In Cohort A, subsets of 3, 5, 7, 9, 12, 15, 20, and 25 experts were randomly sampled without replacement from the complete 27-expert panel. These sizes were selected to represent the range from small to near-full panels rather than to assume an optimal panel size. One thousand random subsets were generated at each panel size.

For each sampled panel, a majority-vote consensus was generated. Agreement with the full-panel consensus, consensus stability, within-panel disagreement, and association with disease progression were assessed.

In Cohort B, the smaller expert pool permitted exhaustive combinatorial analysis.

Odd-numbered panel sizes of 3, 5, 7, 9, and 11 experts were used to reduce tied majority votes and simplify comparisons across panel sizes. For each panel size, all possible expert combinations were enumerated. For each combination, consensus grade was generated by majority vote and its association with survival was quantified using the global penalized likelihood-ratio statistic.

Detailed procedures are provided in Supplementary Methods.

### Expert-composition analysis

For Cohorts A and B, each expert was represented by their classification profile across all cases.

Pairwise Euclidean distances were calculated between expert classification profiles, followed by Ward hierarchical clustering (18). Cluster construction used diagnostic behavior only; clinical outcomes were not used.

Cluster-specific majority-vote consensus classifications were subsequently compared with disease progression in Cohort A and overall survival in Cohort B.

Threshold sensitivity, bootstrap cluster stability (19, 20), and random-expert analyses are described in Supplementary Methods.

### Post hoc expert interpretation of Cohort A and B

After completion of the primary analyses, additional exploratory reviews were conducted to investigate the morphological basis of clinically relevant expert disagreement.

In Cohort A, participating experts reviewed cases in which the expert group showing the strongest observed association with disease progression differed from the overall majority. Particular attention was given to cases classified as UIP by this expert group that subsequently progressed despite a non-UIP full-panel majority. This exercise was hypothesis-generating and was not used to reclassify these cases as UIP.

In Cohort B, selected cases classified as Grade 2 by the all-rater consensus but Grade 3 by Cluster 2 were reviewed to explore morphological features potentially underlying these discordant classifications. These observations were exploratory and did not alter the original grading classifications.

### Statistical analysis

Interobserver agreement was quantified using Krippendorff’s α (21).

In Cohort A, association between consensus UIP classification and disease progression was assessed using Spearman’s rank correlation coefficient (22). Sensitivity and specificity were additionally calculated for binary UIP classification.

In Cohort B, survival associations were evaluated using Firth penalized Cox regression, which reduces estimation bias and improves stability when sample sizes or event counts are limited (23, 24). Overall prognostic separation was quantified using the global penalized likelihood-ratio statistic, which assesses how much better survival outcomes can be explained by the expert-defined groups compared with a model that does not distinguish between these groups. Higher values indicate stronger prognostic separation.

In Cohort C, the association between consensus morphology and disease category was quantified using Cramér’s V, which indicates how closely the observed morphological patterns were related to the underlying disease categories, with higher values indicating a stronger association.

Bootstrap, random-expert, clustering-stability, and threshold-sensitivity procedures are described in Supplementary Methods.

Statistical analyses were performed in Google Colab using Python 3.13.15 with pandas, NumPy, SciPy, and scikit-learn. Firth penalized Cox regression was performed in R using the coxphf package (version 1.13.4).

## Results

### Expert disagreement persisted despite substantial subspecialty experience

The three cohorts comprised internationally distributed pulmonary pathology experts with substantial subspecialty experience (Table 1).

Despite this expertise, interobserver agreement remained incomplete. In Cohort A, Krippendorff’s α was 0.319 for binary UIP classification. In Cohort B, α was 0.501. In Cohort C, α increased from 0.351 before Delphi feedback to 0.539 afterward.

### Larger panels reduced variability but did not increase external clinical relevance

In Cohort A, increasing panel size progressively narrowed the distribution of consensus results across expert compositions.

Median Spearman’s ρ between panel-derived UIP classification and disease progression was 0.174 for three-expert panels and converged toward 0.130 in the near-full and full-panel analyses (Figure 2A).

**Figure 2.**
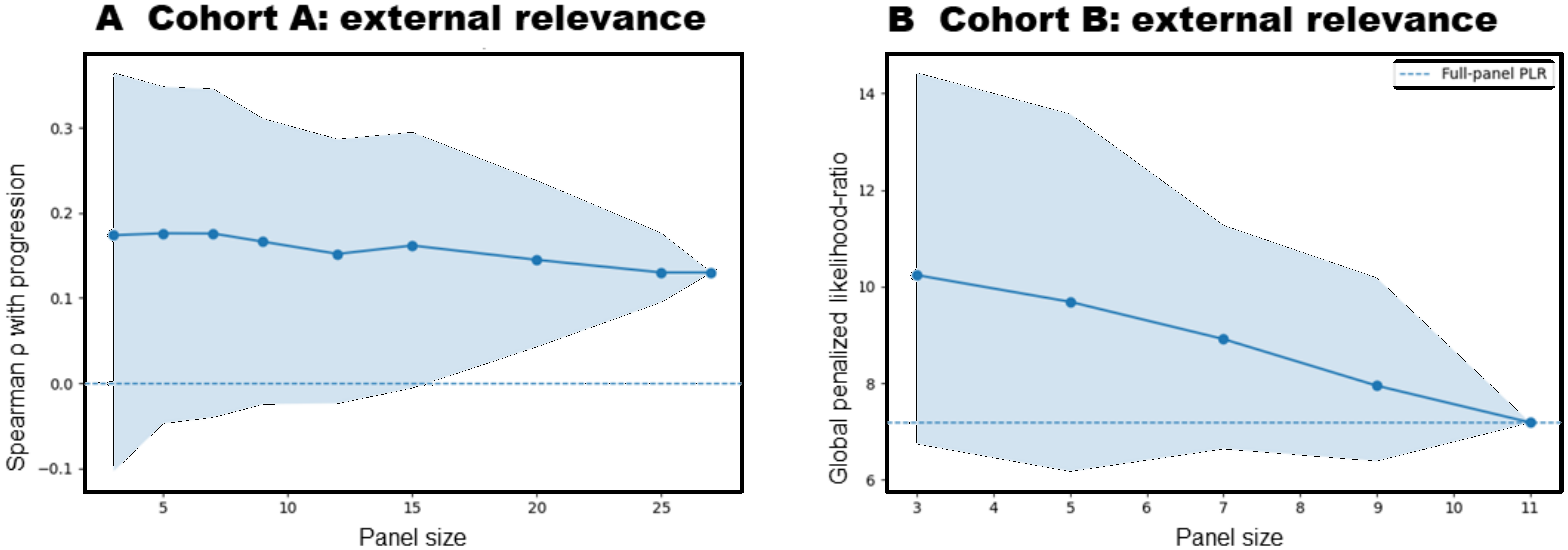
Panel size does not increase external clinical relevance. **A, Cohort A**. Median Spearman’s ρ between panel-derived UIP classification and disease progression across increasing panel sizes. Shading indicates the 2.5th–97.5th percentile range across sampled expert panels. **B, Cohort B**. Median global penalized likelihood-ratio statistic for survival across odd-numbered panel sizes. All possible expert combinations were exhaustively evaluated; shading shows the distribution across panel compositions, and the dashed line indicates the full-panel value. In both cohorts, larger panels reduced variability across expert compositions but did not increase external clinical relevance.

A parallel pattern was observed in Cohort B. Exhaustive analysis of all pathologist combinations at odd-numbered panel sizes showed a progressive decline in median global penalized likelihood-ratio statistics as panel size increased, with narrowing variability and convergence toward the full 11-pathologist panel value (Figure 2B).

Thus, in two independent cohorts using different diseases, pathologist pools, and clinical endpoints, increasing panel size reduced dependence on panel composition but did not increase external clinical relevance.

### Expert composition changed the clinical information represented by consensus

In Cohort A, classification behavior identified three expert groups with different associations between consensus UIP classification and disease progression (Figure 3A).

**Figure 3.**
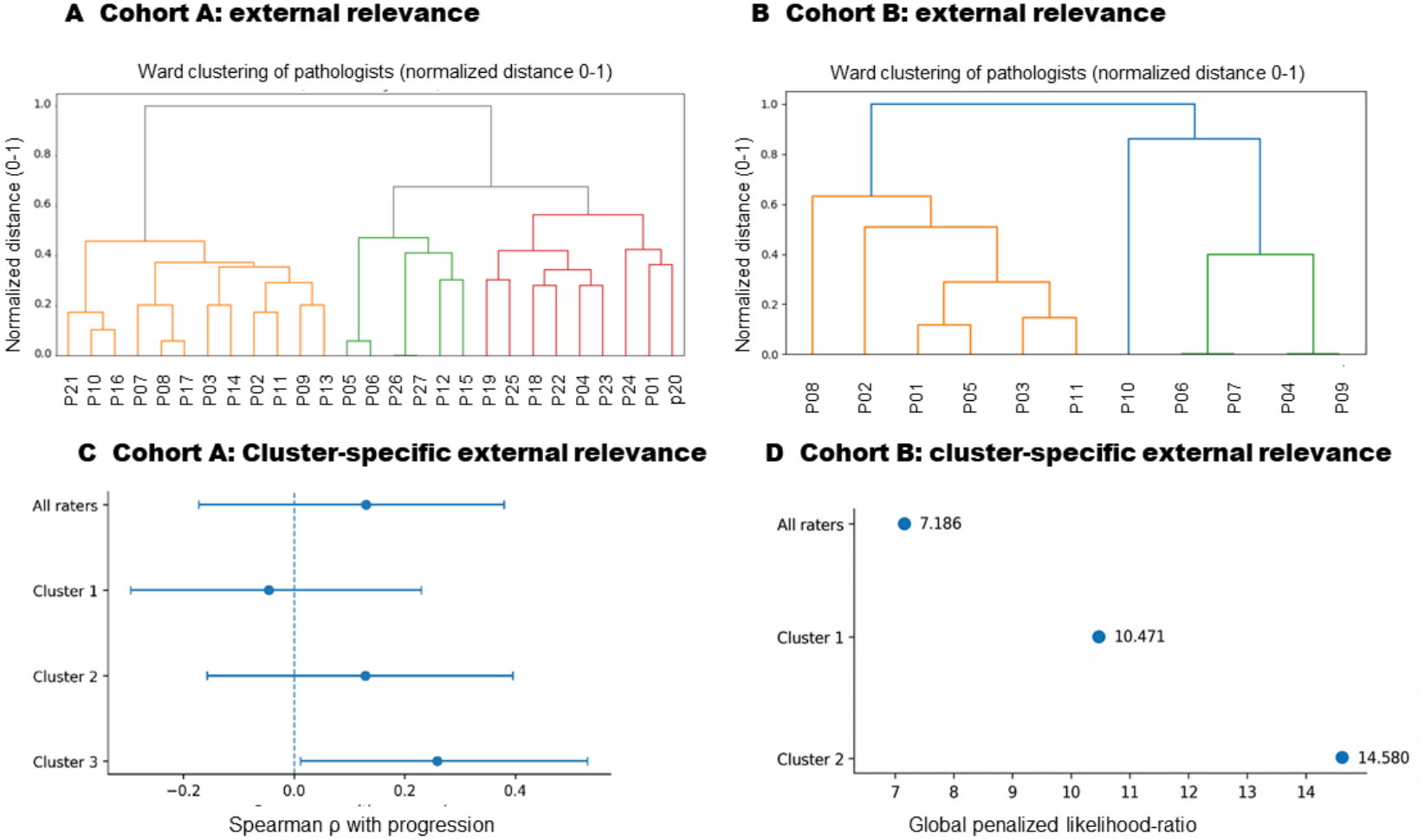
Expert composition changes the clinical information encoded in consensus. **A, B**, Ward hierarchical clustering of experts based solely on classification patterns in Cohorts A and B. Clinical outcomes were not used for cluster formation. **C**, Cluster-specific association between UIP classification and disease progression in Cohort A, expressed as Spearman’s ρ with bootstrap 95% confidence intervals. **D**, Cluster-specific association between histological grade and overall survival in Cohort B, expressed as the global penalized likelihood-ratio statistic.

Spearman’s ρ was −0.046, 0.129, and 0.259 for Clusters 1, 2, and 3, respectively, compared with 0.130 for the full expert panel (Figure 3C).

In Cohort B, pathologist groups identified solely from classification behavior also differed in prognostic association (Figure 3B). The global penalized likelihood-ratio statistic was 7.186 for the full panel, 10.471 for Cluster 1, and 14.580 for Cluster 2 (Figure 3D).

Among the five discordant patients who died, Cluster 2 assigned a higher grade than the full-panel consensus in all cases, including four graded as Grade 2 by the full panel but Grade 3 by Cluster 2. Exploratory review of these four cases identified histologic regions potentially contributing to the discrepant grading, suggesting that inter-group differences may reflect identifiable morphological features (Figure 4).

**Figure 4.**
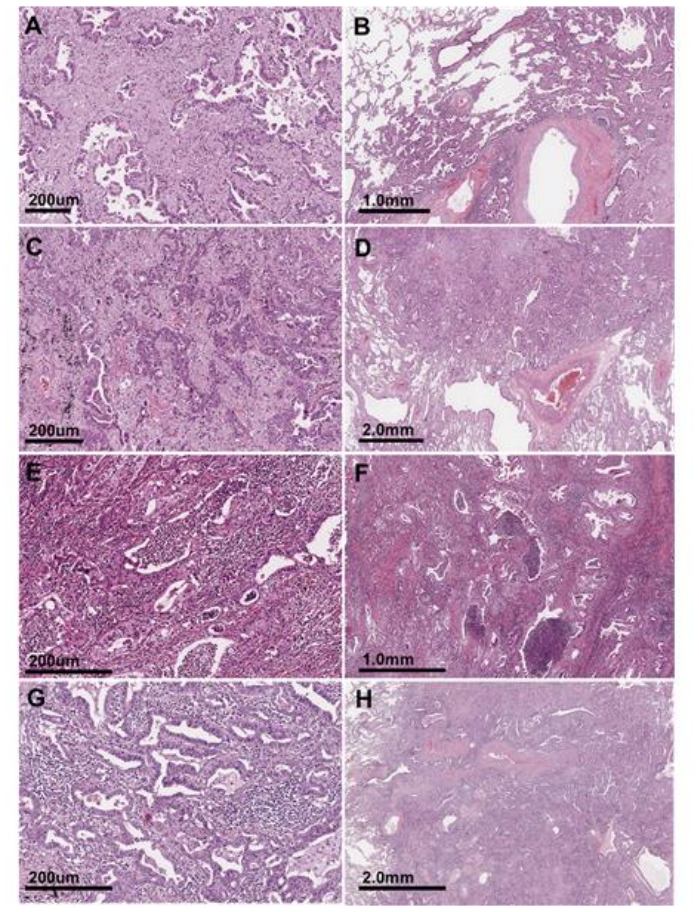
Histologic features underlying discordant grading between the all-rater consensus and Cluster 2. Representative regions from four fatal lung adenocarcinoma cases classified as Grade 2 by the all-rater consensus but as Grade 3 by Cluster 2. These regions illustrate that inter-group grading differences can be localized to specific, extractable morphologic features.

However, random-expert and bootstrap analyses suggested that the observed differences were insufficient to justify defining any expert group as a unique or definitive ground truth (Supplementary Figures S2 and S4).

### Consensus formation improved reproducibility but did not improve external relevance

In Cohort C, modified Delphi feedback increased Krippendorff’s α from 0.351 to 0.539 (Figure 5A).

**Figure 5.**
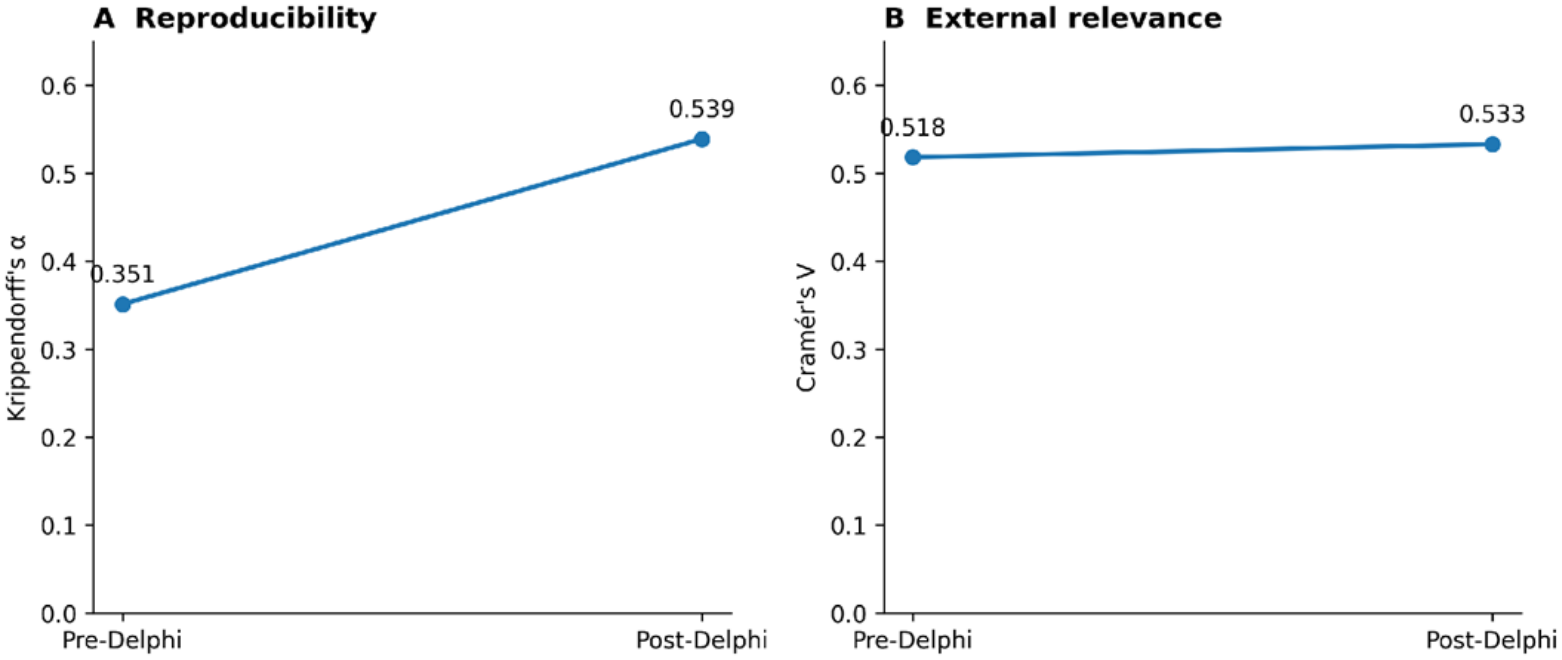
Consensus formation improves reproducibility more than external relevance. **A**, Interobserver reproducibility in Cohort C before and after the modified Delphi process, measured by Krippendorff’s α. **B**, External relevance before and after Delphi consensus, measured by Cramér’s V between consensus morphology and disease category.

By contrast, Cramér’s V between consensus morphology and disease category increased only modestly, from 0.518 to 0.533, with both values indicating a substantial association (Figure 5B).

Thus, structured consensus reduced interobserver variability without a comparable increase in external disease association.

Delphi feedback changed the expert-group structure, but diagnostic differences between groups remained (Supplementary Figures S1 and S3).

## Discussion

This study shows that expert-derived reference standards depend on panel composition, panel size, and how disagreement is resolved.

Across two independent cohorts, increasing panel size reduced variability across expert compositions but did not strengthen association with clinical outcomes. In a third cohort, Delphi consensus improved agreement while only modestly changing disease association. Thus, reproducibility and clinical relevance are distinct properties of an expert-derived reference standard.

These findings extend established consensus methodology, which emphasizes appropriate expert selection, independent judgment, transparency, and structured deliberation (4, 25, 26). Our study addresses a subsequent problem: what should be done when well-qualified experts still disagree?

Increasing panel size should improve the stability of estimates of prevailing expert opinion (27, 28), and our data showed progressively narrower distributions with larger panels. However, disagreement persisted, and clinical association did not improve correspondingly. Larger panels may therefore stabilize majority consensus without resolving the underlying differences in expert interpretation.

This does not imply that smaller panels are superior. Smaller panels were more variable, and some apparent changes in clinical association may reflect convergence toward the full-panel estimate. The key point is that greater consensus stability should not be assumed to indicate greater clinical validity.

Expert composition also mattered. Groups defined without outcome information subsequently showed different associations with progression and survival. This suggests that some expert disagreement may reflect structured differences in interpretation rather than purely random labeling error (11, 14).

This distinction is particularly relevant to AI. Medical-image studies commonly treat disagreement as label noise, although reference labels themselves depend on annotation procedures and panel composition (8, 10, 11). Persistent discordance among expert pathologists has likewise been documented, including in melanocytic lesions, with direct implications for AI ground truth (13, 14, 15).

The solution is not to assign greater authority to the subgroup showing the strongest retrospective association with outcome. Doing so may risk circularity and overfitting the reference standard to the same dataset used to identify that subgroup (29, 30).

Instead, we propose separating expert-signal discovery from reference-standard construction. When one expert group shows greater clinical relevance, the next question should be: What did these experts recognize that others did not? Discordant cases can be examined for candidate morphological features, thresholds, or combinations of findings, which should then be tested in independent cases and, ideally, independent expert populations (16, 31, 32).

Our exploratory review of discordantly graded lung adenocarcinomas illustrates this approach, identifying morphological regions that may help explain differences between expert-group classifications. Although these observations do not establish independent prognostic significance, they provide a starting point for identifying candidate features that could refine existing consensus criteria.

Our previous lung adenocarcinoma studies illustrate one possible downstream application: coherent expert groups were used to generate alternative reference datasets and train distinct deep-learning systems (6, 14).

The same principle may also inform guideline and consensus development, although these processes additionally consider evidence quality, patient values, benefits and harms, and feasibility (33,34). Our data do not establish a new consensus methodology, but suggest that clinically informative disagreement should be characterized before it is eliminated through formal consensus.

Rather than replacing existing consensus standards, independently validated outcome-associated morphological features could be incorporated into these frameworks to improve their clinical relevance while preserving the benefits of expert agreement.

## Limitations

This study has several limitations.

First, expert-panel sizes were not predetermined. Participants were specialists who volunteered for the respective international AI reference-standard projects and therefore do not constitute random samples of pulmonary pathologists.

Second, external clinical relevance does not establish morphological truth. Progression, survival, and disease category were external anchors rather than definitive standards of diagnostic correctness (3, 4).

Third, the three dimensions of reference-standard design were not tested identically across all three cohorts. Panel-size effects were evaluated in Cohorts A and B, whereas structured consensus formation was evaluated directly only in Cohort C.

Fourth, expert clustering was exploratory and showed incomplete stability under resampling. Expert groups should therefore not be interpreted as fixed biological or cognitive phenotypes.

Finally, Cohort C used multiple images from individual patients, and image-level analyses may not fully account for within-patient dependence (35).

## Conclusion

Human consensus should not automatically be equated with clinical ground truth. In expert pathology assessment, greater consensus stability did not necessarily indicate greater clinical relevance. Expert disagreement may contain clinically informative morphological signals that are overlooked by majority consensus. Identifying and independently validating these signals could help refine existing consensus frameworks, moving toward more clinically meaningful reference standards for pathology AI.

## Data Availability

The data supporting the findings of this study are available from the corresponding author upon reasonable request, subject to institutional and ethical restrictions concerning patient confidentiality.

## Supplementary Information

### Supplementary Methods

#### Supplementary Methods 1. Cohort A panel-size analysis

Panel sizes of 3, 5, 7, 9, 12, 15, 20, and 25 experts were evaluated. For each panel size, 1,000 subsets were sampled without replacement from the 27-expert pool, and majority-vote consensus classifications were generated.

Agreement with the full-panel consensus, consensus stability, within-panel disagreement, and Spearman’s ρ with disease progression were calculated. Distributions were summarized using medians and 2.5th–97.5th percentiles.

The full-panel consensus was used as a reference for stability analyses but was not assumed to represent biological ground truth.

#### Supplementary Methods 2. Cohort B exhaustive panel-size analysis

All possible expert combinations were evaluated for odd-numbered panel sizes of 3, 5, 7, 9, and 11 experts.

For each combination, majority-vote consensus grade was generated and association with overall survival was quantified using the global penalized likelihood-ratio statistic.

#### Supplementary Methods 3. Hierarchical clustering of expert judgment

Experts were clustered according to their classifications across cases or images using Euclidean distance and Ward hierarchical clustering. Linkage distances were normalized to 0–1.

Clinical outcomes and disease categories were not used for clustering. Cluster-specific consensus classifications were subsequently evaluated against the relevant external clinical criterion.

#### Supplementary Methods 4. Cluster-threshold sensitivity

Normalized dendrogram thresholds from 0.30 to 0.80 were examined. Cluster membership at each threshold was compared with the primary solution using Jaccard similarity:

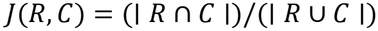

where *R* represents the reference cluster and *C* the corresponding cluster at an alternative threshold (20).

#### Supplementary Methods 5. Random-expert analyses

To assess whether clinically informative observed expert groups could arise from arbitrary expert composition, null analyses were performed.

In Cohort A, random groups containing the same number of experts as the observed high-relevance cluster were repeatedly sampled from the complete expert pool. Consensus classifications and Spearman’s ρ with progression were calculated.

In Cohort B, pathologists were randomly partitioned and the highest global penalized likelihood-ratio statistic within each random partition was retained.

Observed cluster values were compared with the corresponding null distributions.

#### Supplementary Methods 6. Bootstrap analyses

Case-level bootstrap resampling was performed for Cohorts A and B (19).

For Cohort A, 1,000 bootstrap datasets were generated and Spearman’s ρ was recalculated for the full panel and individual expert clusters.

Expert-clustering stability was evaluated using bootstrap resampling followed by repeated hierarchical clustering.

Adjusted Rand indices (36), Jaccard similarities, and pairwise co-clustering probabilities were used to characterize stability.

In Cohort C, bootstrap analyses were conducted at the image level.

#### Supplementary Methods 7. Modified Delphi procedure

After completion of Cohort C Round 1, individual responses were aggregated anonymously.

Participants were shown aggregate classification distributions without disclosure of expert identities. The same experts then independently re-evaluated the images.

Pre- and post-Delphi assessments were compared in terms of Krippendorff’s α, expert-group structure, consensus morphology, and Cramér’s V with disease category.

#### Supplementary Methods 8. Post hoc clinically anchored Cohort A exemplars

Following completion of the primary Cohort A analyses, experts reviewed cases in relation to the observed cluster-specific progression associations.

Three exploratory categories were proposed:

##### Clinically concordant UIP

UIP classification by both the clinically informative expert group and overall majority, with subsequent progression.

##### Outcome-supported discordant UIP

UIP classification by the clinically informative expert group with progression despite non-UIP classification by the overall majority.

##### Clinically concordant non-UIP

non-UIP classification by both the clinically informative group and overall majority with improvement in pulmonary function.

These categories were developed after review of outcome information and were therefore considered hypothesis-generating rather than validated reference labels.

## Supplementary Results

### Supplementary Results 1. Detailed Cohort A panel-size analysis

Increasing panel size progressively increased agreement with the full-panel consensus.

**Supplementary Table S1.**
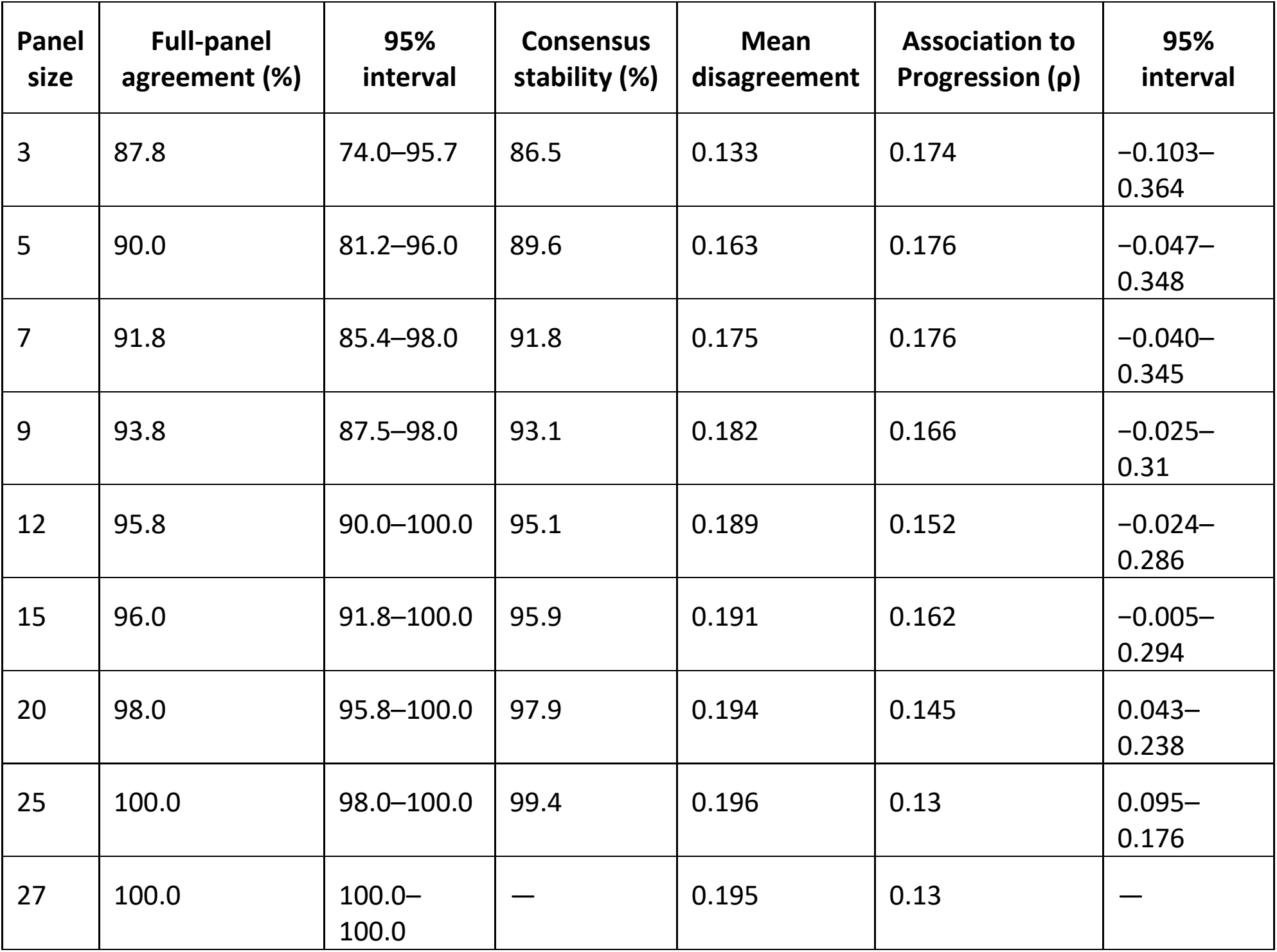

| **Panel size** | **Full-panel agreement (%)** | **95% interval** | **Consensus stability (%)** | **Mean disagreement** | **Association to Progression (ρ)** | **95% interval** |
|---|---|---|---|---|---|---|
| 3 | 87.8 | 74.0–95.7 | 86.5 | 0.133 | 0.174 | −0.103–0.364 |
| 5 | 90.0 | 81.2–96.0 | 89.6 | 0.163 | 0.176 | −0.047–0.348 |
| 7 | 91.8 | 85.4–98.0 | 91.8 | 0.175 | 0.176 | −0.040–0.345 |
| 9 | 93.8 | 87.5–98.0 | 93.1 | 0.182 | 0.166 | −0.025–0.31 |
| 12 | 95.8 | 90.0–100.0 | 95.1 | 0.189 | 0.152 | −0.024–0.286 |
| 15 | 96.0 | 91.8–100.0 | 95.9 | 0.191 | 0.162 | −0.005–0.294 |
| 20 | 98.0 | 95.8–100.0 | 97.9 | 0.194 | 0.145 | 0.043–0.238 |
| 25 | 100.0 | 98.0–100.0 | 99.4 | 0.196 | 0.13 | 0.095–0.176 |
| 27 | 100.0 | 100.0–100.0 | — | 0.195 | 0.13 | — |

Although consensus stability increased progressively, external clinical relevance did not.

### Supplementary Results 2. Cohort B panel-size analysis

All possible expert combinations were evaluated at panel sizes of 3, 5, 7, 9, and 11.

Median global penalized likelihood-ratio statistics declined progressively with increasing panel size and converged toward the full-panel value.

**Supplementary Table S2.**

| Panel size | No. possible combinations | Median global PLR | Distribution interval |
|---|---|---|---|
| 3 | 165 | 10.236 | 6.746-14.417 |
| 5 | 462 | 9.67 | 6.360-13.181 |
| 7 | 330 | 8.912 | 6.392-11.233 |
| 9 | 55 | 7.947 | 6.662-9.695 |
| 11 | 1 | 7.186 | — |

### Supplementary Results 3. Cohort A expert-composition analysis

Hierarchical clustering identified three expert groups.

**Supplementary Table S3.**

| Analysis group | No. experts | Krippendorff’s α | Spearman’s ρ | Bootstrap estimate | 95% CI |
|---|---|---|---|---|---|
| All experts | 27 | 0.319† | 0.130 | 0.127 | −0.172–0.380 |
| Cluster 1 | 12 | 0.280 | −0.046 | −0.050 | −0.295–0.230 |
| Cluster 2 | 6 | 0.556 | 0.129 | 0.129 | −0.157–0.396 |
| Cluster 3 | 9 | 0.422 | 0.259 | 0.256 | 0.012–0.531 |

†Binary UIP classification.

Cluster 3 showed the strongest association with progression, but this did not establish it as a definitive ground truth.

### Supplementary Results 4. Cohort A binary classification

**Supplementary Table S4.**

| Consensus group | Sensitivity, % | Specificity, % |
|---|---|---|
| All experts | 30.4 | 80.8 |
| Cluster 1 | 8.7 | 88.5 |
| Cluster 2 | 42.9 | 73.1 |
| Cluster 3 | 47.8 | 76.9 |

### Supplementary Results 5. Cohort B pathologist-composition analysis

Expert groups showed broadly similar internal agreement but substantially different survival associations.

**Supplementary Table S5.**

| Analysis group | Krippendorff’s α | Global PLR | Global P |
|---|---|---|---|
| All pathologists | 0.501 | 7.186 | 0.02752 |
| Cluster 1 | 0.573 | 10.471 | 0.00532 |
| Cluster 2 | 0.597 | 14.58 | 0.000682 |

Grade-specific Cox estimates were:

**Supplementary Table S6.**

| Group | Comparison | HR | Bootstrap median HR | 95% CI |
|---|---|---|---|---|
| All pathologists | Grade 2 vs 1 | 5.164 | 5.513 | 1.587–23.235 |
|  | Grade 3 vs 1 | 6.404 | 7.131 | 1.825–30.049 |
| Cluster 1 | Grade 2 vs 1 | 5.457 | 6.39 | 1.321–50.103 |
|  | Grade 3 vs 1 | 9.974 | 11.737 | 2.245–93.757 |
| Cluster 2 | Grade 2 vs 1 | 8.556 | 8.556 | 1.129–1169.815 |
|  | Grade 3 vs 1 | 23.743 | 23.743 | 3.244–3126.160 |

### Supplementary Results 6. Random-expert analyses

In Cohort A, randomly constituted expert groups of the same size as Cluster 3 produced a median Spearman’s ρ of approximately 0.163. Approximately 13% generated a value equal to or greater than the observed Cluster 3 value of 0.259 (Figure S2 A).

In Cohort B, random-partition analyses showed that approximately 0.8% of random partitions produced a best-group global PLR statistic equal to or greater than the observed value of 14.580 (Figure S2 B).

These analyses indicate that the clinically informative expert groups were unusual but do not establish them as uniquely optimal.

### Supplementary Results 7. Delphi effects in Cohort C

Krippendorff’s α increased from 0.351 before Delphi consensus to 0.539 afterward. Cramér’s V between morphology and disease category changed from 0.518 to 0.533. Cluster-specific Cramér’s V values were:

**Supplementary Table S7.**

| Assessment | Group | Cramér’s V |
|---|---|---|
| Pre-Delphi | All experts | 0.518 |
|  | Cluster 1 | 0.397 |
|  | Cluster 2 | 0.48 |
| Post-Delphi | All experts | 0.533 |
|  | Cluster 1 | 0.512 |
|  | Cluster 2 | 0.515 |
|  | Cluster 3 | 0.533 |

The range of cluster-specific disease associations narrowed after Delphi consensus.

### Supplementary Results 8. Cluster-threshold sensitivity

Primary expert-group structures persisted across neighboring dendrogram thresholds.

In Cohort A, the principal three-cluster configuration was reproduced at normalized thresholds of approximately 0.60–0.65.

In Cohort B, major reference groups remained stable across neighboring thresholds, including approximately 0.55–0.60.

In Cohort C, the pre-Delphi structure remained relatively stable across a broad intermediate range, whereas the post-Delphi three-group configuration was reproduced most clearly at higher thresholds.

### Supplementary Results 9. Bootstrap cluster stability

Median adjusted Rand indices were approximately:

- Cohort A: 0.364
- Cohort B: 0.416
- Cohort C pre-Delphi: 0.580
- Cohort C post-Delphi: 0.386.

These findings support structured but incompletely stable expert-judgment patterns and argue against interpreting the clusters as fixed expert phenotypes.

## Supplementary Figures and Legends

**Supplementary Figure S1.**
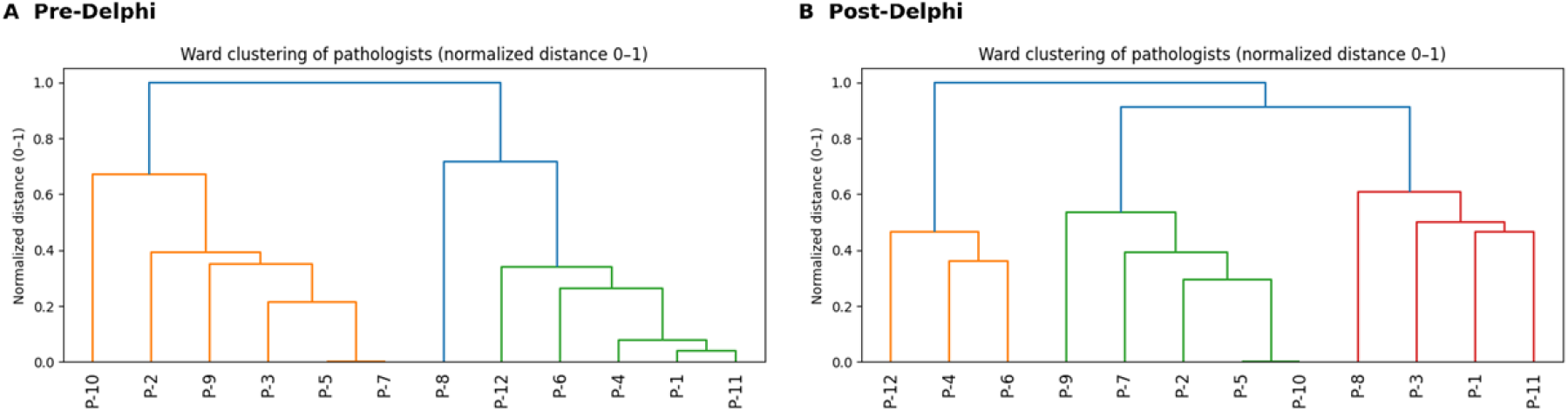
Expert-judgment structure before and after Delphi consensus. Ward hierarchical clustering of Cohort C experts based on granuloma morphology classifications **before (A)** and **after (B)** the modified Delphi process. Distances were normalized to 0–1.

**Supplementary Figure S2.**
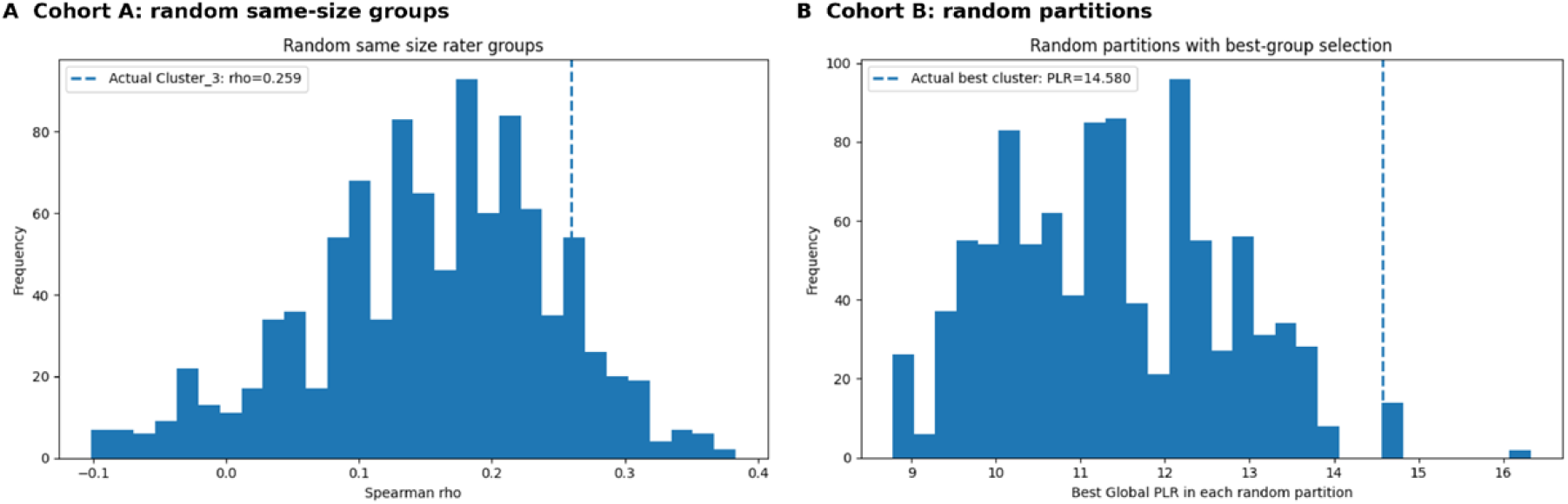
Null analyses for clinically informative expert groups. **A, Cohort A**. Distribution of Spearman’s ρ from randomly constituted expert groups of the same size as the observed high-relevance cluster; dashed line indicates the observed Cluster 3 value. **B, Cohort B**. Distribution of the best global PLR statistic obtained from random pathologist partitions; dashed line indicates the observed best-cluster value.

**Supplementary Figure S3.**
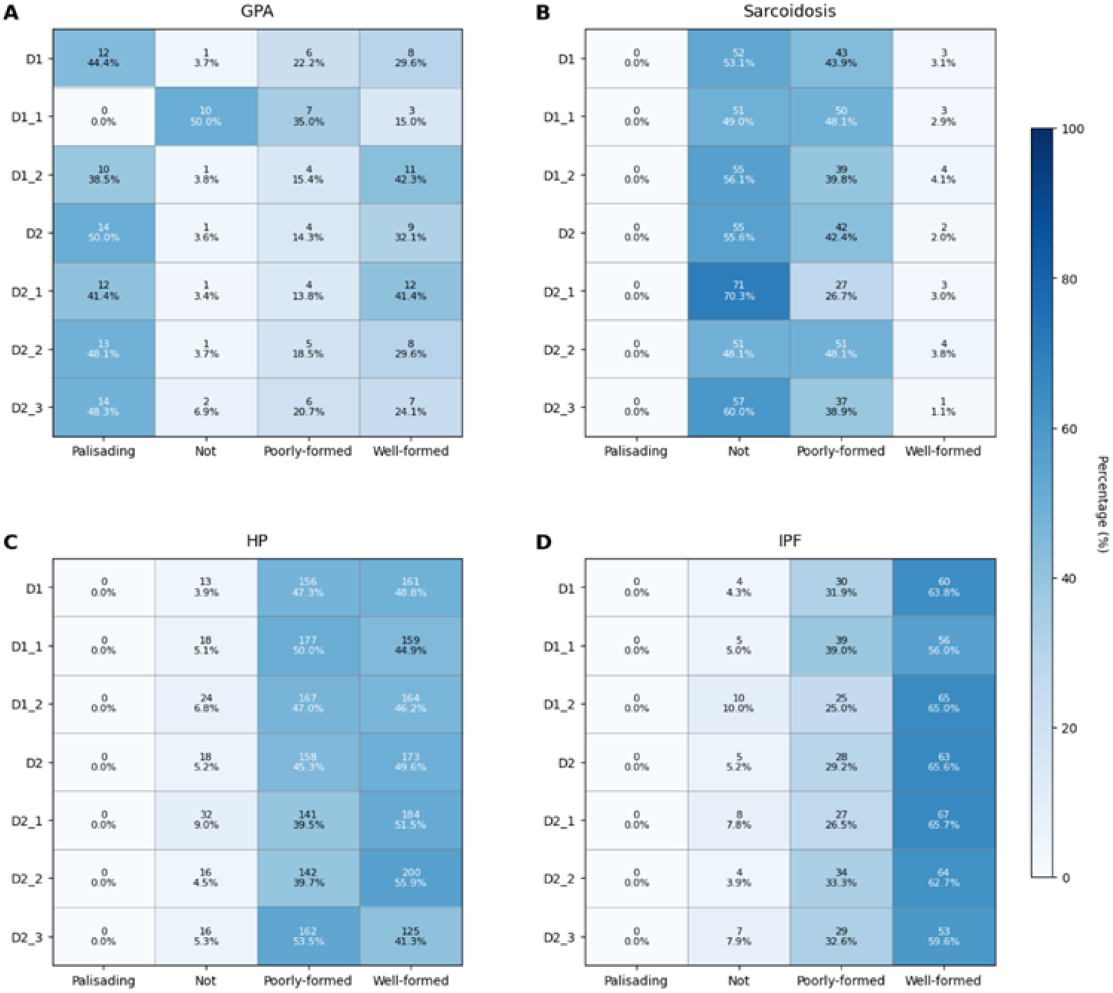
Disease-specific morphology distributions across expert groups. Distribution of consensus granuloma morphologies across GPA, sarcoidosis, hypersensitivity pneumonitis, and IPF in the full expert panel and expert clusters before and after Delphi consensus. Values indicate counts and percentages.

**Supplementary Figure S4.**
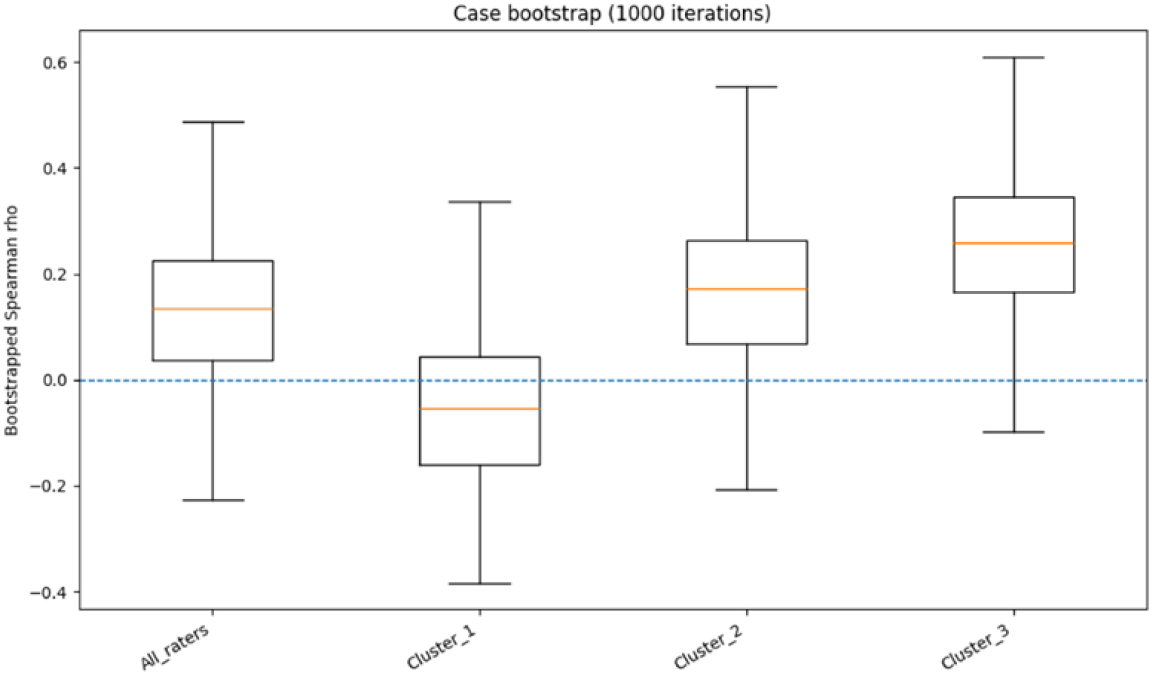
Case-bootstrap distribution of progression association in Cohort A. Distribution of Spearman’s ρ between UIP classification and disease progression across 1,000 case-bootstrap iterations for the full expert panel and each expert cluster.

